# Rehabilitation interventions to modify physical frailty in adults before lung transplantation: A systematic review protocol

**DOI:** 10.1101/2023.08.04.23293669

**Authors:** Laura McGarrigle, Gill Norman, Helen Hurst, Chris Todd

**Affiliations:** Manchester University NHS Foundation Trust; National Institute for Health and Care Research Applied Research Collaboration Greater Manchester; School of Health Sciences, Faculty of Biology, Medicine and Health, The University of Manchester; School of Health and Society, University of Salford; Northern Care Alliance NHS Foundation Trust

## Abstract

**Introduction:** Lung transplantation is the gold standard treatment for end-stage lung disease for a small group of patients meeting strict acceptance criteria after optimal medical management has failed. Physical frailty is prevalent in lung transplant candidates and has been linked to worse outcomes both on the waiting list and postoperatively. Exercise has been proven to be beneficial in optimising exercise capacity and quality of life in lung transplant candidates but its impact on physical frailty is unknown. This review aims to assess the effectiveness of exercise interventions in modifying physical frailty for adults awaiting lung transplantation.

**Methods and Analysis:** This protocol was prospectively registered on the PROSPERO database. We will search 4 databases plus trials registries to identify primary studies including adult candidates for lung transplantation undertaking exercise interventions and assessing outcomes pertaining to physical frailty. Studies must include at least ten participants. Article screening will be performed by two researchers independently at each stage. Extraction will be performed by one reviewer and checked by a second. The risk of bias in studies will be assessed by two independent reviewers using tools appropriate for the research design of each study; where appropriate we will use Cochrane risk of bias 2 or ROBINS-I. At each stage of the review process discrepancies will be resolved through consensus or consultation with a third reviewer. Meta-analyses of frailty outcomes, will be performed if possible and appropriate as will pre-specified subgroup and sensitivity analyses. Where we are unable to perform meta-analysis we will conduct narrative synthesis following SWiM guidance. The review will be reported using the PRISMA Checklist.

**Ethics and Dissemination:** No ethical issues are predicted due to the nature of this study. Dissemination will occur via conference abstracts, professional networks, peer reviewed journals and patient support groups.

**Registration Details:** PROSPERO registration number CRD42022363730.

**Strengths and Limitations of this study:**

- Rigorous systematic review methods at all stages of the review combined with clinical expertise will allow us to produce a reliable first synthesis of the evidence for the effectiveness of rehabilitation in lung transplant candidates for physical frailty.
- A comprehensive search for relevant studies from multiple databases and other sources will allow us to identify relevant studies wherever published. The exclusion of non-English language studies is a limitation of this study but we will list these studies where we identify them.
- We will search for literature pertaining to “lung transplant candidates”, those on the “waiting list” or people “being assessed for lung transplant”. We have developed our search in consultation with an information specialist and it is likely to identify all studies where lung transplant candidates are a substantial proportion of the patient group. However it is not feasible to review every study of individuals with chronic lung disease, particularly where data are unstratified by disease severity or where the authors do not identify transplant candidates. It is therefore possible that we may miss some studies with some relevant data but this is unlikely to substantively impact the review outcomes.
- Using outcomes as a key criterion for inclusion risks missing some relevant studies due to the potential for reporting bias. To mitigate this, we will attempt to contact authors of all otherwise relevant studies to establish if any further outcomes were assessed but not reported and, where possible, obtain relevant data.

## INTRODUCTION

### Lung Transplantation

Lung transplantation (LTx) involves surgically replacing diseased organs in individuals with advanced respiratory failure due to a range of lung diseases including chronic obstructive pulmonary disease (COPD), interstitial lung diseases (ILD), pulmonary arterial hypertension (PAH) and cystic fibrosis (CF). Poor lung function is associated with reduced exercise tolerance, dyspnoea and disability. The terminal stages of these conditions have a profound impact upon individual’s physical function and quality of life. Lung transplantation is a well-established therapy for chronic lung disease in a very specific population who meet the stringent, internationally accepted criteria(1). The strict criteria are necessary to optimise outcomes and protect from inappropriate allocation of such a scare resource. On average, there are around 350 adults on the active lung transplant waiting list in the UK. In the ten years prior to the COVID-19 pandemic, the mean number of lung transplants performed annually in the UK was 178 (±18)(2).

### Frailty

Frailty is a state characterised by lack of physiological reserve and increased vulnerability to stressors and is common in chronic end-stage lung disease (3,4), particularly in those referred for lung transplants (5). There has been a significant rise in the proportion of lung transplant recipients aged over 65 years (6), and increasing age is an independent risk factor for both poor outcomes after LTx (7) and increased incidence of frailty (8).

The two main models conceptualising frailty are the phenotypic and cumulative deficit models. The phenotypic model is more commonly used in LTx and considers frailty through five characteristics; shrinking (weight loss), weakness, exhaustion, slowness and low physical activity(9). The cumulative deficit model conceptualises frailty as an accumulation of symptoms, comorbidities, diseases and health deficiencies. The greater the number of “deficits”, the higher the frailty of the individual (10). Although previously linked to decreased survival after LTx (11), the validity of the latter model in predicting other LTx outcomes is unclear (5).

Although psychosocial, nutritional and physical impairments impact post-LTx outcomes (1), the clinical operationalisation of frailty measurement in LTx relates overwhelmingly to the physical domain (5,12,13). This may be due to the feasibility of data collection, the younger age of LTx candidates or the complexity associated with cumulative deficit models (12,14). The most commonly reported phenotypic frailty measures collected in clinical practice prior to LTx include the Short Physical Performance Battery (SPPB), and the Fried Frailty Phenotype (FFP) (13,15).

Research studies in LTx reflect this phenotypic approach. Although conceptualisation and investigation of biomarkers and their links to frailty (16) (including in patients with lung disease) has recently been undertaken, the field has lacked established markers or cut-offs, preventing their routine use in the evaluation of frailty for LTx (5,12,13). The recent development of a LTx specific frailty scale, the Lung Transplant Frailty Scale (LT-FS) and its validation in 342 LTx candidates may help to resolve this (13). This uses body composition (skeletal muscle mass and percentage body fat), research-grade serum biomarkers and clinical laboratory blood markers in addition to measurement of physical function (SPPB and FFP). All models of the scale displayed superior predictive validity for short term mortality and removal from the LTx waiting list than the FFP or SPPB alone, but long term outcomes have not yet been evaluated.

Previous reviews have identified physical frailty as being detrimental to morbidity and mortality both before and after LTx (17). Physical frailty is also associated with an increased risk of readmission after LTx (18). Transplant teams experience the challenges of identifying patients with the physical and psychological reserve necessary to survive such a demanding peri and postoperative period and to thrive long-term (1,12). This challenge has been exacerbated by the lack of consensus on the optimal assessment tools for frailty in this field (1), which the LT-FS may help to resolve (13). Despite the current lack of consensus on tool utility, it is widely agreed that further work is required to establish the effectiveness of interventions to modify physical frailty, improve candidate selection and LTx outcomes (1,13,19).

### Exercise and Rehabilitation Prior to Lung Transplantation

Rehabilitation for individuals with lung conditions usually takes the form of pulmonary rehabilitation; an evidence-based programme of exercise interventions and education. It aims to reduce dyspnoea, optimise functional capacity, increase participation, and reduce healthcare costs through exacerbations and hospital admissions (20). Rehabilitation whilst on the waiting list (sometimes known as prehabilitation) is recommended for all LTx candidates(1). Rehabilitation after LTx is also a vital component of the recovery programme after surgery and ICU/hospital stay (21). The course of the SARS COVID-19 pandemic has seen many pulmonary rehabilitation schemes develop virtual or telephone offers alongside a face-to-face programme. Technological advances have also led to the development of digital alternatives (22).

### Potential Impact of the Intervention

Advancing lung disease impacts on exercise capacity and accelerates muscle atrophy. Rehabilitation prior to LTx has been shown to improve exercise capacity and quality of life with some evidence of increases in muscle strength (21). Exercise has the potential to contribute to an increase in muscle strength, balance and physical activity which in turn impacts on physical frailty (23). Pulmonary rehabilitation has been shown to reverse frailty in the short term in individuals with COPD when assessed using the FFP (24).

### Why it is Important to do this Review

Professionals in the transplant community are increasingly measuring frailty in attempts to evaluate surgical suitability and weigh the risks and potential benefits of LTx for older individuals. There is emerging evidence linking frailty to reduced quality of life and mortality on the lung transplant waiting list and postoperative adverse outcomes including increased length of hospital stay, readmission, disability and worse health related quality of life (5). Frailty is therefore a significant concern for transplant programmes.

The aim of LTx is to improve survival, function and quality of life of the recipient, whilst considering the ethical elements of who will benefit from such a limited pool of donor organs (12). Understanding and targeting reversible components of frailty has the potential to improve physical condition ahead of major surgery, reduce waiting list mortality and potentially impact post-operative outcomes and maximise benefit from LTx (4,12,25).

Identifying the best strategies to improve frailty prior to transplantation has been highlighted as an area that requires investigation (4,5,13).

Previous systematic reviews have documented the beneficial effect of rehabilitation on exercise capacity and quality of life prior to a lung transplant (21,26). However, there are no prior reviews specifically addressing interventions in modifying physical frailty in this population despite the significance of the problem and link to poor outcomes (17).

### Aims and Objectives

This systematic review aims to evaluate the impact of exercise interventions in modifying physical frailty for adults awaiting lung transplantation. We also aim to identify any harms that occur as a result of an exercise intervention.

## METHODS AND ANALYSIS

### Study Design

This study will be a systematic review with methodology following guidance from the Cochrane Handbook of Systematic Reviews (27). It will be reported in accordance with the Preferred Reporting Items for Systematic Reviews and Meta-Analyses checklist (PRISMA) (28). Any important amendments will be documented on Prospero.

### Inclusion and Exclusion Criteria

#### Types of Studies

Preliminary searches indicated a very few randomised controlled trials (RCT) and non-randomised controlled studies, which we initially planned to include. We therefore widened the inclusion criteria to original primary research studies with any design, including those without controls, with more than 10 participants; this amendment was registered on Prospero. We therefore anticipate including randomised and non-randomised trials, cohort studies and case series. Cross-sectional studies may be eligible if they assess frailty following an intervention. We will not include evidence syntheses, commentaries, case studies, and non-systematic narrative reviews.

#### Types of Participants

We will include studies of adults over the age of 18, on a waiting list (candidates) for a single or double lung transplant. Individuals with any underlying lung disease who have met the criteria to be accepted onto the waiting list at a lung transplant centre, where frailty has been assessed will be included. There are well established and accepted internationally agreed criteria for lung transplantation (1). We will include studies involving candidates for primary LTx or subsequent repeat LTx procedures. LTx performed by any surgical incision site will be included. Studies of candidates listed for a multi-organ transplant in the same surgical procedure (including but not limited to, concurrent heart and lung or lung and liver) will be excluded unless data is provided for lung recipients only or multi-organ candidates comprise under 25% of study We will exclude studies of people completing interventions whilst receiving extra corporeal membrane oxygenation (ECMO) as a bridge to transplant; they are restricted to interventions within the intensive care unit which is not the focus of this review.

#### Types of Intervention

We will include any formal physical exercise or physical activity delivered prescribed under professional guidance which includes formal evaluation of outcomes. The intervention may be supervised or unsupervised, face to face or virtual and performed in any setting (community or hospital). There is no minimum length or intensity of intervention. The intervention may be single (one form of exercise) or multimodal (for example a combination of strength and endurance training). The intervention must contain an exercise or physical activity component. Physical activity is defined as any bodily movement produced by skeletal muscles that results in energy expenditure. Exercise is defined as a subset of physical activity that is planned, structured and repetitive and has an objective of the improvement or maintenance of physical fitness (29).

#### Types of Comparators

We will accept any comparator or no comparator. Where studies include comparison groups we anticipate identifying comparisons of rehabilitation with one or more of the following: no intervention, “usual care”, advice only, or an alternative intervention which may or may not meet our intervention criteria. The specific rehabilitation intervention should be the only systematic difference between the groups.

#### Outcomes

We will consider the following physical frailty measures as primary outcomes: measures of Phenotypic frailty (FFP) or cumulative deficit frailty models (e.g. Clinical Frailty Scale, Electronic Frailty Index,).

We will also include surrogate markers of frailty e.g. SPPB, sarcopenia via hand grip dynamometry, quadriceps force, CT scan (muscle cross sectional area), muscle strength testing of upper of lower limb: manual or non-manual (e.g. 1 rep max), sit to stand testing and objective assessments of balance.

Studies must include at least one direct or indirect measure of frailty in order to be eligible for inclusion. Where no relevant outcome is reported we will attempt to contact authors of all otherwise relevant studies to establish if any further outcomes were assessed but not reported and, where possible, obtain relevant data. We will distinguish between studies excluded after confirmation that no relevant outcomes were assessed and those where this confirmation was not possible.

Where studies report a relevant primary outcome we will consider the following as secondary outcomes: Mortality (on waiting list or post-operatively), hospital/ICU length of stay, health-related quality of life measures. Any adverse events reported during interventions will be recorded.

### Search Strategies

Literature search strategies have been developed using medical subject headings (MeSH) related to rehabilitation, exercise and lung transplant candidates with support from an experienced medical librarian. Trials will be identified from searches of the following databases: MEDLINE (Ovid) 1980 to date, EMBASE (Ovid) 1980 to date, CINHAL Plus (EBSCO) 1980 to date, Cochrane Central Register of Controlled Trials (CENTRAL), the Cochrane Library, trials registries (ClinicalTrials.gov and the WHO trials portal). All databases will be searched from 1980 to the present. The success of lung transplantation was established only after the discovery and introduction of the immunosuppressive agent cyclosporine which became accepted practice in the early 1980s (30). A draft search strategy for MEDLINE is given in Table 1 with this strategy being adopted and modified for other databases (see supplementary material).

**Table 1.**
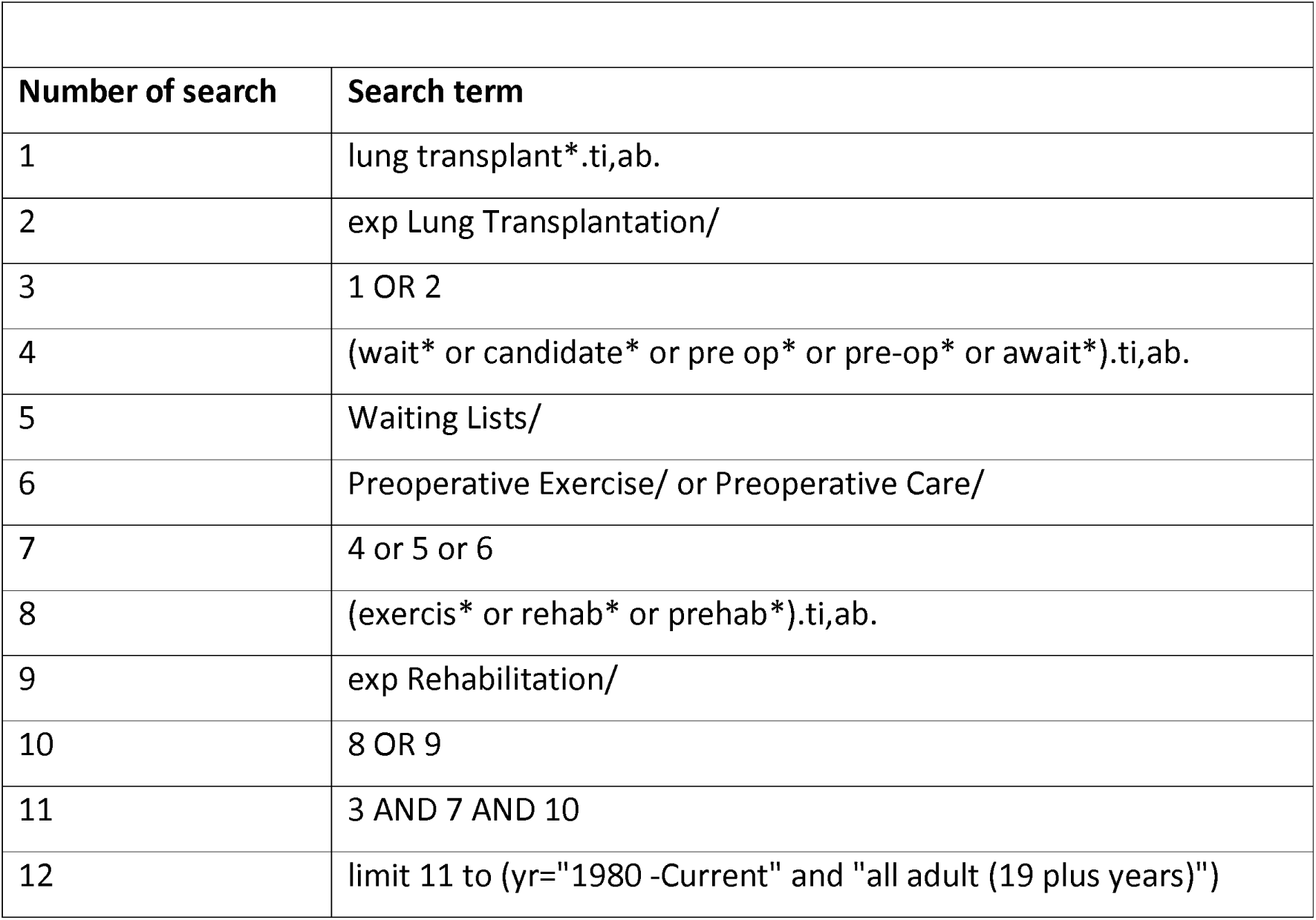
Ovid MEDLINE(R) Search strategy.

| Number of search | Search term |
| --- | --- |
| 1 | lung transplant*.ti,ab. |
| 2 | exp Lung Transplantation/ |
| 3 | 1 OR 2 |
| 4 | (wait* or candidate* or pre op* or pre-op* or await*).ti,ab. |
| 5 | Waiting Lists/ |
| 6 | Preoperative Exercise/ or Preoperative Care/ |
| 7 | 4 or 5 or 6 |
| 8 | (exercis* or rehab* or prehab*).ti,ab. |
| 9 | exp Rehabilitation/ |
| 10 | 8 OR 9 |
| 11 | 3 AND 7 AND 10 |
| 12 | limit 11 to (yr="1980 -Current" and "all adult (19 plus years)") |

We will also search the references of all identified included studies and of identified systematic or scoping reviews.

### Selecting Studies

Following deduplication in Endnote, title and abstract and full text screening will be performed by two independent reviewers (LM and GN) against the pre-defined inclusion criteria using Rayyan software (31). Any discrepancies between reviewers will be resolved by consensus or consultation with a third reviewer (HH or CT). Any non-English language studies will be retained and listed for reference but not included in the synthesis process. This will enhance transparency, mitigate language publication bias and maps the extent of missing evidence in our results.

### Data Extraction and Management

Two authors will pilot and agree a data extraction form. Data extraction will be completed by one author (LM) and checked by a second (GN). Any discrepancy will be reconciled by consensus or a third author (HH or CT). The data extraction form will be based on the information collated in Table 2.

**Table 2:** Data extraction plan.

| **Table 2: Data extraction plan.** |  |
| --- | --- |
|  | **Data to be extracted** |
| **Study Design** | Design (e.g. RCT)<br>Date of study period<br>Country and setting e.g. single site or multi-centre study<br>Duration of follow-up<br>Sample size calculation<br>Ethical approval and (methods of) informed consent |
| **Participants** | Inclusion/Exclusion Criteria<br>Characteristics for each group including:<br>Number of participants<br>Age (Mean (s.d.))<br>Ratio of male:female<br>Disease/diagnosis<br>Co-morbidities<br>Primary transplant (or repeat transplant after graft failure); number/percentage per group.<br>Characteristics that stratify health opportunities and outcomes (PROGRESS-plus) (32) |
| **Interventions** | Based on TIDieR: Template for Intervention Description and Replication Checklist (33).<br>Type of exercise component/other intervention in each group<br>Frequency, intensity and duration of intervention (including how this was prescribed)<br>Location of intervention delivery (e.g. Home, hospital, community, gym) |
|  | Who delivered the intervention<br>Mode of intervention delivery (e.g. Face to face, telephone, video call)<br>Tailoring / modification<br>Adherence measured? |
| **Outcomes and measures of effect** | Definition for each outcome alongside minimal clinically important difference (MCID) where available, scale limits and direction of benefit.<br>Time points measured.<br>Validation of tool<br><u>Outcomes:</u> For each group and each outcome at each time point we will extract the number of participants with data in each group and the following data:<br>Continuous outcomes: mean with s.d. for each group. Medians with IQR will be extracted where reported. Mean differences or standardised mean differences with 95% CI will be extracted where these are the only reported data. P values will be extracted in the absence of other outcome data as will descriptive reporting of results.<br>Dichotomous outcomes: number in each group with event. For adverse events types of adverse events will also be reported. Relative risks or odds ratios with 95% CI will be extracted where these are the only reported data.<br>Time to event data: hazard ratios or data to calculate these. |
| **Other** | Other relevant details of study design or intervention.<br>Funding Source<br>Conflicts of interest<br>Recruitment failure<br>Patient and public involvement or engagement |

Where outcomes use continuous scales of measurement (e.g. composite frailty scores, muscle cross-sectional area) we will express the results as the mean difference or, where scales or units are unclear or composites, standardised mean difference with 95% confidence intervals (CI). We will express dichotomous outcomes as risk ratios (RR) with 95% CI. Survival outcomes will be reported as hazard ratios with 95% CI. If outcomes are reported at multiple time points we will extract all the data, but will analyse primary data at the end of the intervention and at the latest time point reported.

### Missing Data

Any missing data will be requested from the original author by email and included in the review. If we are unable to gather the missing data, where possible, we will calculate or impute these (e.g. where measures of variance are not available). Where outcome data remain missing and cannot be imputed we will use assumptions in analysing dichotomous data; that is we will assume that missing participants did not have the outcome assessed but we will explore the impact of this assumption with a sensitivity analysis. For continuous outcomes we will perform a completed case analysis and will not attempt to impute results for missing participants.

### Risk of Bias Assessment

RCTs and quasi-RCTs will be assessed using the Cochrane Risk of Bias tool (RoB2) with an extension for cluster or crossover RCTs as required (27). ROBINS-I (34) or another appropriate tool will be used for non-randomised controlled studies. Other study designs (e.g. cohort and cross-sectional studies and case-series) will be assessed using an appropriate tool. Risk of bias assessments will be done by two reviewers independently and any discrepancies reconciled by a 3 ^rd^ reviewer.

### Data analysis - Strategy for Data Synthesis

Clinical and methodological heterogeneity will be considered initially. If the studies identified vary widely in terms of population, study design, intervention and outcomes a meta-analysis may be inappropriate and therefore a narrative synthesis of findings will be produced and reported following the Synthesis without Meta-analysis (SWiM) guidelines for narrative evidence synthesis (35). If sufficient studies are identified with low heterogeneity, statistical heterogeneity will be considered to determine if a random-effects meta-analysis can be performed.

### Assessment of heterogeneity

We will initially assess statistical heterogeneity by visual inspection of the forest plot followed by a quantification using the I ^2^ statistic. (75% - 100% considerable heterogeneity; 50-90% considered to represent substantial heterogeneity; 30-60% moderate and 0-40% considered unimportant heterogeneity) (27)

### Subgroup analysis

Participant heterogeneity may be investigated using subgroup analysis by disease type (COPD, CF, IPF for example). Subgroup analysis by age may be considered (18-69, and ≥70 years) as lung transplant recipients over the age of 70 years have a decreased longer term survival (36). Intervention heterogeneity may be investigated by subgroup analysis including the following potential groups: virtual and in person rehabilitation, exercise intensity, exercise type (e.g. aerobic versus resistance) or the degree of supervision (e.g. fully supervised versus non supervised). We will exercise caution in implementing these prespecified analyses and are mindful of the risks of performing multiple such analyses.

### Sensitivity analysis

If appropriate we will use sensitivity analyses to explore the impact of any imputation of or assumptions about missing data. If we have performed a meta-analysis with very few studies we will conduct a fixed effects analysis to assess the impact of the random effects model in this context (37).

### Summary of Findings tables

We will present a summary of the main findings of the review in a table format including key information regarding the magnitude of effects for each outcome alongside a rating of the certainty of evidence. We will use the Grading of Recommendations Assessment, Development and Evaluation working group methodology considerations (risk of bias, consistency of effect, imprecision, indirectness and publication bias) to assess the quality of the body of evidence for each prespecified outcome (38,39). We will justify all decisions to down- or up-grade the certainty of the evidence. If appropriate we will present the evidence in summary of findings tables generated using GRADEpro software (40).

### Patient and Public Involvement

Local lung transplant support group attendees have been involved in some preliminary patient and public engagement work prior to the data collection for this study. Further PPI work will be completed to contribute to the production of a plain language summary and insight into dissemination strategies.

## ETHICS AND DISSEMINATION

This systematic review has been registered prospectively (registration number CRD42022363730). Its findings will be submitted to international transplantation conferences and peer reviewed journals for dissemination of the results and conclusions.

Results, and their implications will be shared at local and national meetings of LTx clinicians.

We will also share findings with people living with chronic lung disease on the transplant waiting list and their carers via local patient support groups.

## Data Availability

This is a protocol for a systematic review and there is therefore no data linked to this manuscript at present.

## ACKNOWLEDGEMENTS AND STATEMENTS OF FINANCIAL SUPPORT

LM holds a part-time predoctoral fellowship funded by the National Institute for Health and Care Research Applied Research Collaboration Greater Manchester (NIHR ARC-GM)(Grant award number NIHR200174). CT is a CI and partially funded and GN is wholly funded by NIHR ARC-GM. The views expressed in this publication are those of the authors and not necessarily those of the National Institute for Health and Care Research or the Department of Health and Social Care or its partner organisations.

## Contributions

LM had the idea for the review, wrote the first draft of the protocol and designed the search strategy; GN, HH and CT advised on the protocol, edited and commented substantively on the protocol; all authors approved the manuscript prior to submission.

## Acknowledgements

The authors thank Bethan Morgan (librarian at Manchester University NHS Foundation Trust) for her support with electronic search strategies for this review.

## REFERENCE LIST

1. Leard LE, Holm AM, Valapour M, et al. Consensus document for the selection of lung transplant candidates: An update from the International Society for Heart and Lung Transplantation. J Heart Lung Transplant. 2021;40(11):1349–79.

2. NHSBT Annual report of cardiothoracic organ transplantation [website] London (UK) 2020 [Updated 2020, cited March 2023]. Available from https://nhsbtdbe.blob.core.windows.net/umbraco-assets-corp/19874/nhsbt-annual-report-on-cardiothoracic-organ-transplantation-201920.pdf

3. Venado A, McCulloch C, Greenland JR, et al. Frailty trajectories in adult lung transplantation: A cohort study. The Journal of Heart and Lung Transplantation. 2019;38(7):699–707.

4. Kobashigawa J, Dadhania D, Bhorade S, et al. Report from the American Society of Transplantation on frailty in solid organ transplantation. American Journal of Transplantation. 2019;19(4):984–94.

5. Varughese R, Rozenberg D, Singer LG. An update on frailty in lung transplantation. Current Opinion in Organ Transplantation. 2020;25(3):274–9.

6. Valapour M, Lehr CJ, Skeans MA, et al. OPTN/SRTR 2018 Annual Data Report: Lung. American Journal of Transplantation. 2020;20(s1):427–508.

7. Courtwright A, Cantu E. Lung transplantation in elderly patients. J Thorac Dis. 2017;9(9):3346–51

8. Clegg A, Young J, Iliffe S, et al. Frailty in elderly people. Lancet. 2013;381(9868):752–62.

9. Fried LP, Tangen CM, Walston J, et al. Frailty in older adults: evidence for a phenotype. J Gerontol A Biol Sci Med Sci. 2001;56(3):M146–56.

10. Rockwood K, Mitnitski A. Frailty in Relation to the Accumulation of Deficits. The Journals of Gerontology: Series A. 2007;62(7):722–7.

11. Wilson ME, Vakil AP, Kandel P, et al. Pretransplant frailty is associated with decreased survival after lung transplantation. J Heart Lung Transplant. 2016;35(2):173–8.

12. Agarwal A, Neujahr DC. Frailty in lung transplantation: Candidate assessment and optimization. Transplantation. 2021;105(10):2201–12.

13. Singer JP, Christie JD, Diamond JM, et al. Development of the Lung Transplant Frailty Scale (LT-FS). J Heart Lung Transplant. 2023;42(7):892–904.

14. Spiers GF, Kunonga TP, Hall A, et al. Measuring frailty in younger populations: a rapid review of evidence. BMJ Open. 2021;11(3):e047051.

15. Snyder LD, Singer LG. Beyond the eyeball test: Measures of frailty in lung transplantation. The Journal of Heart and Lung Transplantation. 2019;38(7):708–9.

16. Hakeem FF, Maharani A, Todd C, et al. Development, validation and performance of laboratory frailty indices: A scoping review. Archives of Gerontology and Geriatrics. 2023;111:104995.

17. Montgomery E, Macdonald PS, Newton PJ, et al. Frailty in lung transplantation: a systematic review. Expert Review of Respiratory Medicine. 2020;14(2):219–27.

18. Courtwright AM, Zaleski D, Gardo L, et al. Causes, Preventability, and Cost of Unplanned Rehospitalizations Within 30 Days of Discharge After Lung Transplantation. Transplantation. 2018;102(5):838–44

19. Wilson ME, Vakil AP, Kandel P, et al. Pretransplant frailty is associated with decreased survival after lung transplantation. J Heart Lung Transplant. 2016;35(2):173–8.

20. Holland AE, Cox NS, Houchen-Wolloff L, et al. Defining Modern Pulmonary Rehabilitation. An Official American Thoracic Society Workshop Report. Ann Am Thorac Soc. 2021;18(5):e12–e29.

21. Hume E, Ward L, Wilkinson M, et al. Exercise training for lung transplant candidates and recipients: A systematic review. European Respiratory Review. 2020;29(158):1–19.

22. Singer JP, Soong A, Bruun A, et al. A mobile health technology enabled home-based intervention to treat frailty in adult lung transplant candidates: A pilot study. Clin Transplant. 2018;32(6):e13274.

23. Angulo J, El Assar M, Álvarez-Bustos A, et al. Physical activity and exercise: Strategies to manage frailty. Redox Biol. 2020;35:101513.

24. Maddocks M, Kon SSC, Canavan JL, et al. Physical frailty and pulmonary rehabilitation in COPD: a prospective cohort study. Thorax. 2016;71(11):988–95.

25. Rozenberg D, Orsso CE, Chohan K, et al. Clinical outcomes associated with computed tomography-based body composition measures in lung transplantation: a systematic review. Transpl Int. 2020;33(12):1610–25.

26. Hoffman M, Chaves G, Ribeiro-Samora GA, et al. Effects of pulmonary rehabilitation in lung transplant candidates: a systematic review. BMJ open. 2017 Feb 1;7(2):e013445.

27. Higgins JPT, Thomas J, Chandler J, et al. (editors)[website]. Cochrane Handbook for Systematic Reviews of Interventions version 6.3 (updated February 2022). Cochrane, 2022. Available from www.training.cochrane.org/handbook

28. Page MJ, McKenzie JE, Bossuyt PM, et al. The PRISMA 2020 statement: an updated guideline for reporting systematic reviews. BMJ. 2021;372:n71.

29. Casperson CJ, Powell KE, Christenson GM. Physical activity, exercise and physical fitness: Definitions and distinctions for health-related research. Public Health Reports. 1985;100(2):126–31.

30. Venuta F, Van Raemdonck D. History of lung transplantation. J Thorac Dis. 2017;9(12):5458–71.

31. Ouzzani M, Hammady H, Fedorowicz Z, et al. Rayyan—a web and mobile app for systematic reviews. Systematic Reviews. 2016;5(1):210.

32. O’Neill J, Tabish H, Welch V, et al. Applying an equity lens to interventions: using PROGRESS ensures consideration of socially stratifying factors to illuminate inequities in health. Journal of Clinical Epidemiology. 2014;67(1):56–64.

33. Hoffmann TC, Glasziou PP, Boutron I, et al. Better reporting of interventions: template for intervention description and replication (TIDieR) checklist and guide. BMJ : British Medical Journal. 2014;348:g1687.

34. Sterne JAC, Hernán MA, Reeves BC, et al. ROBINS-I: a tool for assessing risk of bias in non-randomised studies of interventions. BMJ. 2016;355:i4919.

35. Campbell M, McKenzie JE, Sowden A, et al. Synthesis without meta-analysis (SWiM) in systematic reviews: reporting guideline. BMJ. 2020;368:l6890.

36. Hayanga AJ, Aboagye JK, Hayanga HE, et al. Contemporary analysis of early outcomes after lung transplantation in the elderly using a national registry. J Heart Lung Transplant. 2015;34(2):182–8.

37. Bender R, Friede T, Koch A, et al. Methods for evidence synthesis in the case of very few studies. Research Synthesis Methods. 2018;9(3):382–92.

38. Guyatt G, Oxman AD, Akl EA, et al. GRADE guidelines: 1. Introduction—GRADE evidence profiles and summary of findings tables. Journal of Clinical Epidemiology. 2011;64(4):383–94.

39. Schünemann H, Brożek J, Guyatt G, Oxman A. GRADE handbook for grading quality of evidence and strength of recommendations. guidelinedevelopment.org/handboo2013

40. GRADEpro GDT: GRADEpro Guideline Development Tool [Software]. McMaster University and Evidence Prime, 2022. Available from gradepro.org

